# Accelerometer-Derived “Weekend Warrior” Physical Activity and Cardiovascular Outcomes Across the Stages 0–3 of Cardiovascular-Kidney-Metabolic Syndrome: A Prospective UK Biobank Study

**DOI:** 10.64898/2026.01.13.26344014

**Authors:** Yuemei Xi, Qiang Wang, Bin Zhu

## Abstract

**Background:** Cardiovascular-Kidney-Metabolic (CKM) syndrome is a progressive spectrum where patients often struggle with daily exercise. The “Weekend Warrior” (WW) pattern offers a flexible alternative, but its efficacy across CKM stages remains unclear. We evaluated the association of accelerometer-derived WW activity with cardiovascular outcomes across CKM Stages 0–3.

**Methods:** This prospective study included 88,832 UK Biobank participants (CKM Stages 0–3). Physical activity was objectively measured via 7-day accelerometry and categorized by weekly moderate-to-vigorous physical activity (MVPA) volume and pattern: Inactive (<150 min), WW (≥150 min; ≥50% volume in 1–2 days), or Regularly Active (RA). The primary outcome was incident cardiovascular disease (CVD), a composite of coronary heart disease (CHD), heart failure (HF), stroke, atrial fibrillation (AF), and peripheral artery disease (PAD).

**Results:** During a median 7.5-year follow-up, 9,125 incident CVD events were recorded. In fully adjusted models, both WW and RA patterns were associated with similar risk reductions for total CVD (WW: HR 0.87, 95% CI 0.83–0.91, P < 0.001; RA: HR 0.88, 95% CI 0.83–0.94, P < 0.001) and CHD (WW: HR 0.86, 95% CI 0.80– 0.93, P < 0.001; RA: HR 0.86, 95% CI 0.79–0.93, P < 0.001). Notably, the WW pattern demonstrated unique benefits for AF (HR 0.91, 95% CI 0.85–0.99, P = 0.024) and PAD (HR 0.85, 95% CI 0.78–0.93, P < 0.001). In CKM Stage 3, the WW pattern showed a 38% reduction in total CVD risk (HR 0.62, 95% CI 0.47–0.83, P = 0.001) and marked reductions in HF (HR 0.47, 95% CI 0.24–0.90, P = 0.022) and PAD (HR 0.52, 95% CI 0.31–0.89, P = 0.019). Dose-response analysis revealed a non-linear L-shaped association, with CVD risk reductions plateauing at 200–300 min/week of MVPA. Furthermore, the WW pattern significantly offset the risk conferred by high genetic susceptibility and elevated inflammation (Stage 3 with hs-CRP > 3 mg/L: HR 0.46, 95% CI 0.25–0.84, P = 0.012). Both patterns conferred substantial survival advantages, with the WW pattern showing a 24% lower risk of all-cause mortality (HR 0.76, 95% CI 0.70–0.81, P < 0.001).

**Conclusions:** The WW pattern is associated with significant reductions in CVD risk and mortality across the CKM spectrum. Concentrating activity into 1–2 days is a feasible, safe, and effective strategy, offering unique vascular protection in advanced disease. For CKM patients, achieving total MVPA volume—ideally 200–300 min/week—should be prioritized over frequency.

## Introduction

The conceptualization of Cardiovascular-Kidney-Metabolic (CKM) syndrome represents a major paradigm shift in chronic disease management. The recent Presidential Advisory from the American Heart Association (AHA) integrates metabolic risk factors, chronic kidney disease (CKD), and the cardiovascular system into a systemic disease spectrum, highlighting the severe challenge it poses to global public health [1,2]. CKM syndrome not only multiplies the risk of premature mortality and major adverse cardiovascular events (MACE) but is also accelerating due to the concurrent epidemics of obesity and diabetes [3]. Given the complex multi-organ crosstalk inherent to CKM syndrome, there is an urgent need to identify universal intervention strategies that span the entire disease continuum—from early metabolic risk (Stage 0–1) to advanced clinical disease (Stage 3).

Physical activity (PA) is universally recognized as a cornerstone lifestyle intervention in the comprehensive management of cardiovascular disease. However, while guidelines recommend at least 150 minutes of moderate-to-vigorous physical activity (MVPA) per week [4], a significant adherence paradox exists in clinical practice: The CKM patients in greatest need of physical activity are frequently facing barriers to maintaining daily exercise routines due to multimorbidity, severe fatigue, or occupational time constraints [5]. In this context, the “Weekend Warrior” (WW) pattern—compressing the recommended exercise volume into 1–2 days—has emerged as an attractive alternative due to its high temporal flexibility. Although early questionnaire-based studies suggested benefits [6], and recent large-scale accelerometer-based studies have confirmed the cardiovascular protection of the WW pattern in the general population [7], whether this conclusion applies to the distinct, high-risk CKM population remains unresolved.

Unlike the general healthy population, CKM syndrome constitutes a progressive pathophysiological continuum. As individuals transition from Stage 0 to 3, they experience a progressively accumulating burden of chronic inflammation, endothelial dysfunction, and impaired metabolic flexibility [8,9]. Within this context, it remains unclear whether concentrated, high-load exercise (the WW pattern) continues to confer cardiovascular protection or instead precipitate additional risk. Moreover, it is unclear whether such intermittent high-intensity activity can improve metabolic health to the same extent as regular, sustained physical activity. These knowledge gaps are particularly salient among individuals with advanced CKM syndrome (Stage 3) and those with a dual high-risk profile characterized by both high genetic susceptibility and elevated inflammatory burden. Consequently, there is an urgent need to comprehensively examine the complex interactions between different CKM stages and patterns of physical activity.

Leveraging objective accelerometer data from over 100,000 participants in the UK Biobank prospective cohort, this study aims to: (1) define the association of WW and regularly active (RA) patterns with the risk of cardiovascular disease and its subtypes across the CKM Stages 0–3; (2) integrate polygenic risk scores (PRS) to explore whether physical activity can offset the superimposed risks of congenital genetics and disease progression; and (3) elucidate the potential role of systemic inflammation in this association. Collectively, this study seeks to provide evidence for prescribing precise, personalized physical activity regimens for patients with CKM syndrome—particularly those unable to adhere to regular daily exercise.

## Methods

### 2.1 Study Design and Participants

This study was nested within the UK Biobank, a prospective, population-based cohort comprising over 500,000 individuals aged 37–73 years at recruitment [10]. a sub-cohort of 103,568 participants accepted the invitation to wear a wrist-worn triaxial accelerometer (AX3, Axivity) for seven consecutive days to objectively quantify physical activity. To ensure data integrity, we applied standard UK Biobank quality control protocols. Participants were excluded if their device data could not be successfully calibrated or if the record provided less than 72 hours of valid wear time. Following these exclusions, 100,018 participants with high-quality accelerometer data remained.

The study population was defined according to the CKM syndrome framework proposed by the 2023 American Heart Association (AHA) Presidential Advisory [1]. Participants were categorized into CKM Stages 0 through 4. Given our focus on the role of physical activity in preventing the progression of cardiovascular disease (CVD), individuals with prevalent CVD at the time of accelerometer wear (CKM Stage 4, n = 11,186) were excluded from the analysis. This resulted in a final analytical sample of 88,832 participants across CKM Stages 0 to 3 (Fig. S1). All participants provided electronic informed consent, and the study received ethical approval from the North West Multi-centre Research Ethics Committee (REC reference: 21/NW/0157) and the UK National Information Governance Board.

### 2.2 Physical Activity Assessment and Classification

Physical activity was objectively captured using a triaxial accelerometer (Axivity AX3, Newcastle upon Tyne, UK) worn on the dominant wrist for a continuous seven-day period [11]. The devices recorded raw acceleration at 100 Hz within a ±8g range. Following standard calibration to local gravity and noise filtering, non-wear periods were defined as stationary episodes (acceleration SD <13.0 mg) lasting at least 60 minutes. To quantify activity intensities, a previously validated machine-learning framework that integrates a Random Forest classifier for initial behavior labeling and a Hidden Markov Model (HMM) for temporal smoothing was utilized [12]. This approach enhances classification accuracy by correcting biologically implausible transitions between 30-second epochs. MVPA was defined as any waking behavior reaching an intensity of ≥3 metabolic equivalents. Daily total MVPA volume (Field ID 40045) was aggregated from these classified windows without requiring a minimum bout duration, aligning with current physical activity guidelines [13–15].

Based on World Health Organization (WHO) physical activity guidelines, participants were categorized into three mutually exclusive physical activity patterns: Inactive: total weekly MVPA < 150 minutes. WW: total weekly MVPA ≥ 150 minutes, with ≥ 50% of total MVPA was accumulated within 1–2 days; RA: total weekly MVPA ≥ 150 minutes, with activity distributed more evenly across the week and not meeting the concentration criterion for the WW pattern.

### 2.3. Definition of CKM Syndrome Stages

We staged all participants according to the 2023 AHA guidelines, integrating physical examination, biochemical markers, medication history, and disease diagnoses. A hierarchical classification approach was applied from the highest risk (Stage 3) to the lowest (Stage 0). Detailed definitions, data cleaning procedures, and algorithms for risk prediction are provided in Table S1. Briefly, Stage 3 is defined as high predicted 10-year cardiovascular risk (≥ 20%) calculated using the AHA PREVENT equations [16], or very-high-risk chronic kidney disease (CKD). Stage 2 is defined as the presence of metabolic risk factors (hypertension, type 2 diabetes, or dyslipidemia) or moderate-risk CKD. Stage 1 is defined as overweight/obesity (BMI ≥ 25 kg/m² for non-Asians; ≥ 23 kg/m² for Asians), abdominal obesity, or prediabetes, in the absence of Stage 2 or 3 criteria. Stage 0 is defined as individuals with optimal metabolic profiles and no risk factors.

### 2.4 Covariates

Covariates included sociodemographic characteristics, lifestyle factors, anthropometric measurements, and clinical history. Baseline data were primarily obtained through touchscreen questionnaires, nurse-led interviews, and physical examinations. Sociodemographic variables included age, sex, ethnicity, educational attainment, and the Townsend Deprivation Index (TDI). Lifestyle factors included smoking status (never, previous, current) and alcohol consumption (never, previous, current). Diet quality was assessed using the Healthy Diet Score (HDS), calculated based on the frequency of intake of fruits, vegetables, fish, processed meat, and red meat [17]. Sleep quality was evaluated using a composite Sleep Score incorporating sleep duration, insomnia symptoms, and snoring[18]. Body mass index (BMI) was calculated as weight (kg) divided by height squared (m²). Estimated glomerular filtration rate based on creatinine and cystatin C (eGFR*cr*–*cy*s) was calculated using the CKD-EPI 2021 equation [19]. A history of hypertension, diabetes, and dyslipidemia was defined by self-reported physician diagnosis, relevant medication use, or corresponding hospital inpatient records. Medication use, including antihypertensive, lipid-lowering, and glucose-lowering therapies, was ascertained by combining specific medication records from nurse-led interviews with broad medication categories reported in touchscreen questionnaires. Detailed definitions, UK Biobank Field IDs, and categorization of covariates are provided in Table S2. Specific medication codes are listed in Table S3.

### 2.5 Assessment of Systemic Inflammation and Genetic Susceptibility

To evaluate whether the associations between physical activity and cardiovascular outcomes differed across biological risk profiles, we incorporated markers of systemic inflammation and genetic susceptibility in stratified analyses. Serum hs-CRP levels were measured at baseline using an immunoturbidimetric assay (Beckman Coulter AU5800). In accordance with established AHA and Centers for Disease Control and Prevention (CDC) guidelines, as well as evidence from large UK Biobank studies [20,21], participants were stratified by systemic inflammatory status into elevated inflammation (hs-CRP > 3 mg/L) and low-to-moderate inflammation (hs-CRP ≤ 3 mg/L) groups. This threshold has demonstrated strong clinical relevance for identifying individuals at elevated cardiovascular risk. We used the standardized CVD polygenic risk score provided by UK Biobank (Category 301). This score was derived from large-scale genome-wide association studies (GWAS) using Bayesian regression with continuous shrinkage (LDPred2). To account for potential population stratification, the first 10 genetic principal components (PC1–10) were included as covariates in all genetic analyses.

### 2.6 Ascertainment of CVD

The primary outcome was incident CVD, defined as a composite endpoint comprising CHD, heart failure (HF), stroke, AF, and PAD. Outcome events were ascertained using UK Biobank’s algorithmically defined “First Occurrence of Health Outcomes” data fields, which integrate data from hospital inpatient records (Hospital Episode Statistics for England, Scotland, and Wales) and national death registries (NHS Digital). These fields report the date of the first record of a 3-character ICD-10 category, encompassing the specific sub-codes detailed in Table S4. The follow-up period commenced on the date of accelerometer attachment and continued until the earliest of the following: the first outcome event, death, loss to follow-up, or the censoring date of October 31, 2022.

### 2.7. Statistical Analysis

Cox proportional hazards regression models were used to estimate the associations between physical activity patterns and the risk of incident CVD, with results expressed as hazard ratios (HRs) and 95% confidence intervals (CIs). To distinguish the total effect of physical activity on cardiovascular health from its independent effect, three sequential models were constructed. Model 1 adjusted for age, sex, and ethnicity. Model 2 further adjusted for socioeconomic status (TDI and educational attainment), lifestyle factors (smoking status, alcohol consumption, HDS, and Sleep Score), and baseline kidney function (eGFR*cr-cys*), capturing the overall effect of physical activity including pathways mediated through metabolic health in accordance with a recent large-scale cohort study [7]. Model 3 (fully adjusted model) additionally adjusted for BMI, histories of hypertension, diabetes, and dyslipidemia, as well as corresponding medication use, and was considered the primary model for CKM stage–specific analyses.

Restricted cubic spline models with four knots placed at the 5th, 35th, 65th, and 95th percentiles were used to assess dose–response relationships between MVPA and CVD risk. To minimize the influence of extreme values on the tails of the spline curves, continuous MVPA values were winsorized at the 95th percentile prior to modeling. For analyses assessing the joint association of physical activity and genetic susceptibility, the PRS was categorized into low/intermediate risk (first and second tertiles) and high risk (third tertile), with additionally adjustment for the first 10 genetic principal components. Stratified analyses were conducted across CKM Stages 0–3, and interaction effects were evaluated using likelihood ratio tests. To assess the robustness of our findings, we conducted a series of sensitivity analyses for the primary outcome (Total CVD): (1) varying the definition of the active group using alternative MVPA volume thresholds (75 min [approx. 25th percentile], median [215 min], and 75th percentile [381 min]); (2) applying a stricter WW criterion requiring ≥75% of total MVPA within 1–2 days; (3) excluding participants diagnosed with CVD within the first 2 years of follow-up to minimize reverse causality; and (4) employing the Fine-Gray subdistribution hazard model to account for the competing risk of all-cause mortality. Furthermore, standard Cox proportional hazards models were utilized to estimate the association between physical activity patterns and all-cause mortality to evaluate overall survival benefit. Additionally, we performed stratified analyses across major demographic and clinical subgroups including age, sex, BMI, TDI, smoking status, alcohol consumption, hypertension, and diabetes status, to assess the consistency of associations. All statistical analyses were performed using R software (version 4.4.0). A two-sided P value < 0.05 was considered statistically significant.

## 3. Results

### 3.1 Baseline Characteristics

The study population included 88,832 participants with CKM syndrome (Stages 0 to 3). Among them, 37,377 (42.1%) followed the WW pattern, making it the most prevalent physical activity pattern in this cohort, followed by the Inactive group (32,661; 36.8%) and the RA group (18,794; 21.2%). Baseline characteristics stratified by physical activity pattern are shown in Table 1. Compared with the two active groups, inactive participants were slightly older (mean age 56.1 years) and had a higher proportion of women (68.0%). Overall, the Inactive group exhibited the most unfavorable demographic and clinical profile. Regarding metabolic health, the Inactive group had a significantly higher mean BMI (27.8 kg/m²) compared with the WW (26.0 kg/m²) and RA (25.7 kg/m²) groups. Consistent with this, the prevalence of major comorbidities was highest in the Inactive group, including hypertension (74.3%), dyslipidemia (61.7%), and diabetes (5.4%).

**Table 1.**
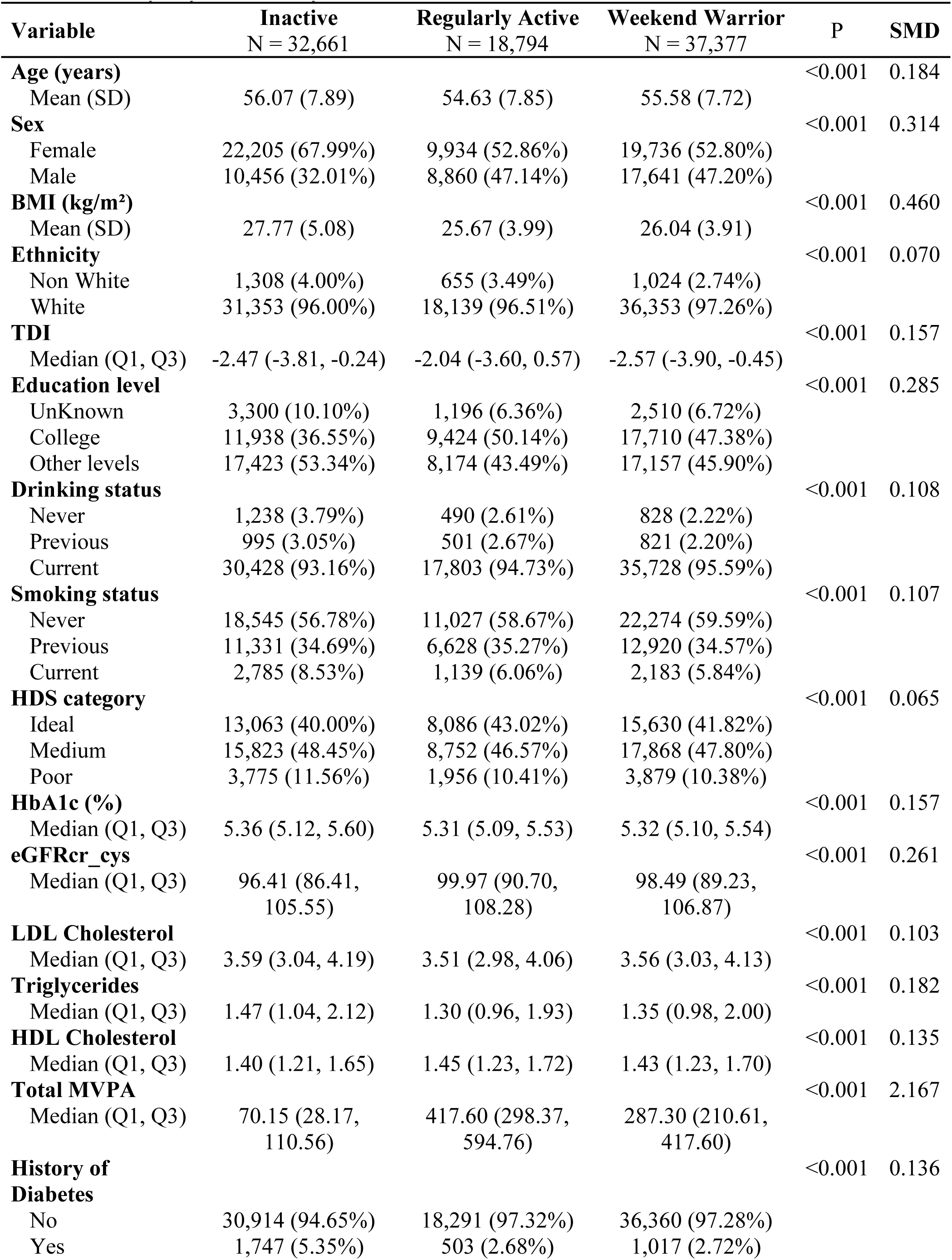
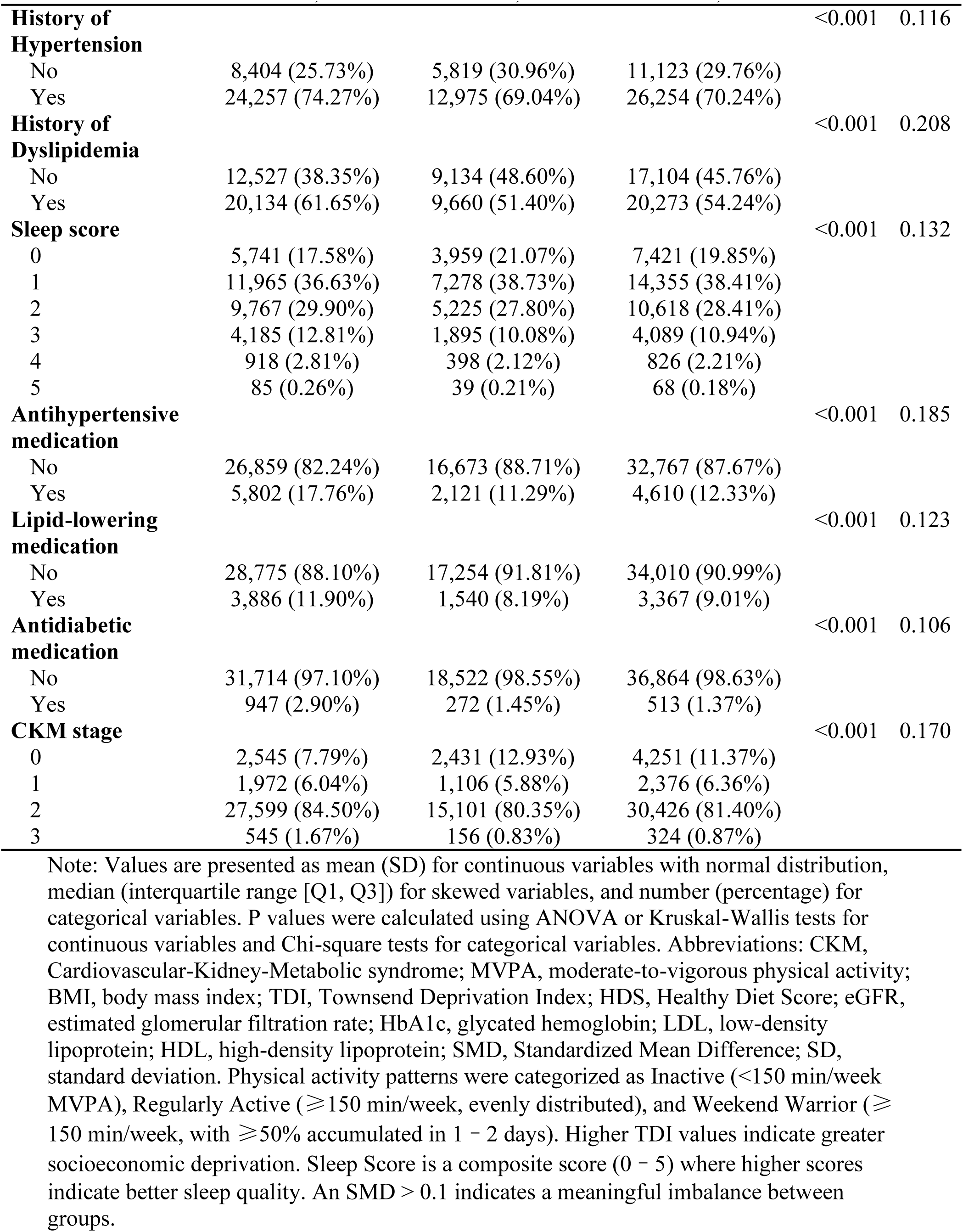
Baseline Characteristics of 88,832 Participants with CKM Syndrome (Stages 0–3) Stratified by Physical Activity Pattern.

| Variable | Inactive<br>N = 32,661 | Regularly Active<br>N = 18,794 | Weekend Warrior<br>N = 37,377 | P | SMD |
| --- | --- | --- | --- | --- | --- |
| **Age (years)** |  |  |  | <0.001 | 0.184 |
| Mean (SD) | 56.07 (7.89) | 54.63 (7.85) | 55.58 (7.72) |  |  |
| **Sex** |  |  |  | <0.001 | 0.314 |
| Female | 22,205 (67.99%) | 9,934 (52.86%) | 19,736 (52.80%) |  |  |
| Male | 10,456 (32.01%) | 8,860 (47.14%) | 17,641 (47.20%) |  |  |
| **BMI (kg/m<sup>2</sup>)** |  |  |  | <0.001 | 0.460 |
| Mean (SD) | 27.77 (5.08) | 25.67 (3.99) | 26.04 (3.91) |  |  |
| **Ethnicity** |  |  |  | <0.001 | 0.070 |
| Non White | 1,308 (4.00%) | 655 (3.49%) | 1,024 (2.74%) |  |  |
| White | 31,353 (96.00%) | 18,139 (96.51%) | 36,353 (97.26%) |  |  |
| **TDI** |  |  |  | <0.001 | 0.157 |
| Median (Q1, Q3) | -2.47 (-3.81, -0.24) | -2.04 (-3.60, 0.57) | -2.57 (-3.90, -0.45) |  |  |
| **Education level** |  |  |  | <0.001 | 0.285 |
| UnKnown | 3,300 (10.10%) | 1,196 (6.36%) | 2,510 (6.72%) |  |  |
| College | 11,938 (36.55%) | 9,424 (50.14%) | 17,710 (47.38%) |  |  |
| Other levels | 17,423 (53.34%) | 8,174 (43.49%) | 17,157 (45.90%) |  |  |
| **Drinking status** |  |  |  | <0.001 | 0.108 |
| Never | 1,238 (3.79%) | 490 (2.61%) | 828 (2.22%) |  |  |
| Previous | 995 (3.05%) | 501 (2.67%) | 821 (2.20%) |  |  |
| Current | 30,428 (93.16%) | 17,803 (94.73%) | 35,728 (95.59%) |  |  |
| **Smoking status** |  |  |  | <0.001 | 0.107 |
| Never | 18,545 (56.78%) | 11,027 (58.67%) | 22,274 (59.59%) |  |  |
| Previous | 11,331 (34.69%) | 6,628 (35.27%) | 12,920 (34.57%) |  |  |
| Current | 2,785 (8.53%) | 1,139 (6.06%) | 2,183 (5.84%) |  |  |
| **HDS category** |  |  |  | <0.001 | 0.065 |
| Ideal | 13,063 (40.00%) | 8,086 (43.02%) | 15,630 (41.82%) |  |  |
| Medium | 15,823 (48.45%) | 8,752 (46.57%) | 17,868 (47.80%) |  |  |
| Poor | 3,775 (11.56%) | 1,956 (10.41%) | 3,879 (10.38%) |  |  |
| **HbA1c (%)** |  |  |  | <0.001 | 0.157 |
| Median (Q1, Q3) | 5.36 (5.12, 5.60) | 5.31 (5.09, 5.53) | 5.32 (5.10, 5.54) |  |  |
| **eGFR<sub>cr</sub>_cys** |  |  |  | <0.001 | 0.261 |
| Median (Q1, Q3) | 96.41 (86.41, 105.55) | 99.97 (90.70, 108.28) | 98.49 (89.23, 106.87) |  |  |
| **LDL Cholesterol** |  |  |  | <0.001 | 0.103 |
| Median (Q1, Q3) | 3.59 (3.04, 4.19) | 3.51 (2.98, 4.06) | 3.56 (3.03, 4.13) |  |  |
| **Triglycerides** |  |  |  | <0.001 | 0.182 |
| Median (Q1, Q3) | 1.47 (1.04, 2.12) | 1.30 (0.96, 1.93) | 1.35 (0.98, 2.00) |  |  |
| **HDL Cholesterol** |  |  |  | <0.001 | 0.135 |
| Median (Q1, Q3) | 1.40 (1.21, 1.65) | 1.45 (1.23, 1.72) | 1.43 (1.23, 1.70) |  |  |
| **Total MVPA** |  |  |  | <0.001 | 2.167 |
| Median (Q1, Q3) | 70.15 (28.17, 110.56) | 417.60 (298.37, 594.76) | 287.30 (210.61, 417.60) |  |  |
| **History of Diabetes** |  |  |  | <0.001 | 0.136 |
| No | 30,914 (94.65%) | 18,291 (97.32%) | 36,360 (97.28%) |  |  |
| Yes | 1,747 (5.35%) | 503 (2.68%) | 1,017 (2.72%) |  |  |
| **History of Hypertension** |  |  |  | <0.001 | 0.116 |
| No | 8,404 (25.73%) | 5,819 (30.96%) | 11,123 (29.76%) |  |  |
| Yes | 24,257 (74.27%) | 12,975 (69.04%) | 26,254 (70.24%) |  |  |
| **History of Dyslipidemia** |  |  |  | <0.001 | 0.208 |
| No | 12,527 (38.35%) | 9,134 (48.60%) | 17,104 (45.76%) |  |  |
| Yes | 20,134 (61.65%) | 9,660 (51.40%) | 20,273 (54.24%) |  |  |
| **Sleep score** |  |  |  | <0.001 | 0.132 |
| 0 | 5,741 (17.58%) | 3,959 (21.07%) | 7,421 (19.85%) |  |  |
| 1 | 11,965 (36.63%) | 7,278 (38.73%) | 14,355 (38.41%) |  |  |
| 2 | 9,767 (29.90%) | 5,225 (27.80%) | 10,618 (28.41%) |  |  |
| 3 | 4,185 (12.81%) | 1,895 (10.08%) | 4,089 (10.94%) |  |  |
| 4 | 918 (2.81%) | 398 (2.12%) | 826 (2.21%) |  |  |
| 5 | 85 (0.26%) | 39 (0.21%) | 68 (0.18%) |  |  |
| **Antihypertensive medication** |  |  |  | <0.001 | 0.185 |
| No | 26,859 (82.24%) | 16,673 (88.71%) | 32,767 (87.67%) |  |  |
| Yes | 5,802 (17.76%) | 2,121 (11.29%) | 4,610 (12.33%) |  |  |
| **Lipid-lowering medication** |  |  |  | <0.001 | 0.123 |
| No | 28,775 (88.10%) | 17,254 (91.81%) | 34,010 (90.99%) |  |  |
| Yes | 3,886 (11.90%) | 1,540 (8.19%) | 3,367 (9.01%) |  |  |
| **Antidiabetic medication** |  |  |  | <0.001 | 0.106 |
| No | 31,714 (97.10%) | 18,522 (98.55%) | 36,864 (98.63%) |  |  |
| Yes | 947 (2.90%) | 272 (1.45%) | 513 (1.37%) |  |  |
| **CKM stage** |  |  |  | <0.001 | 0.170 |
| 0 | 2,545 (7.79%) | 2,431 (12.93%) | 4,251 (11.37%) |  |  |
| 1 | 1,972 (6.04%) | 1,106 (5.88%) | 2,376 (6.36%) |  |  |
| 2 | 27,599 (84.50%) | 15,101 (80.35%) | 30,426 (81.40%) |  |  |
| 3 | 545 (1.67%) | 156 (0.83%) | 324 (0.87%) |  |  |

Note: Values are presented as mean (SD) for continuous variables with normal distribution, median (interquartile range [Q1, Q3]) for skewed variables, and number (percentage) for categorical variables. P values were calculated using ANOVA or Kruskal-Wallis tests for continuous variables and Chi-square tests for categorical variables. Abbreviations: CKM, Cardiovascular-Kidney-Metabolic syndrome; MVPA, moderate-to-vigorous physical activity; BMI, body mass index; TDI, Townsend Deprivation Index; HDS, Healthy Diet Score; eGFR, estimated glomerular filtration rate; HbA1c, glycated hemoglobin; LDL, low-density lipoprotein; HDL, high-density lipoprotein; SMD, Standardized Mean Difference; SD, standard deviation. Physical activity patterns were categorized as Inactive (<150 min/week MVPA), Regularly Active ( ≥ 150 min/week, evenly distributed), and Weekend Warrior ( ≥ 150 min/week, with ≥ 50% accumulated in 1 – 2 days). Higher TDI values indicate greater socioeconomic deprivation. Sleep Score is a composite score (0 – 5) where higher scores indicate better sleep quality. An SMD > 0.1 indicates a meaningful imbalance between groups.

Notably, the WW and RA groups shared similar clinical characteristics. For instance, differences in BMI, diabetes and hypertension rates were minimal. Given the meaningful imbalances in these baseline characteristics (Standardized Mean Differences > 0.1 for many variables), we adjusted for these CKM-related confounders in subsequent multivariable analyses to isolate the independent effects of physical activity.

### 3.2 Associations of Physical Activity with Cardiovascular Outcomes in Individuals with CKM Syndrome (Stages 0–3)

During a median follow-up of 7.5 years, a total of 9,125 incident CVD events were recorded. Figure 1 illustrates the cumulative incidence of total CVD and its subtypes stratified by physical activity patterns among participants across CKM Stages 0 to 3. Kaplan-Meier estimates visually demonstrated that the Inactive group had the highest cumulative risk for all evaluated outcomes (Log-rank test P < 0.001). While the survival curves for the WW and RA groups closely overlapped for most outcomes, the WW pattern showed a trend toward lower cumulative risks for HF and PAD compared with the RA group, suggesting potential additional benefits in these specific phenotypes.

**Figure 1.**
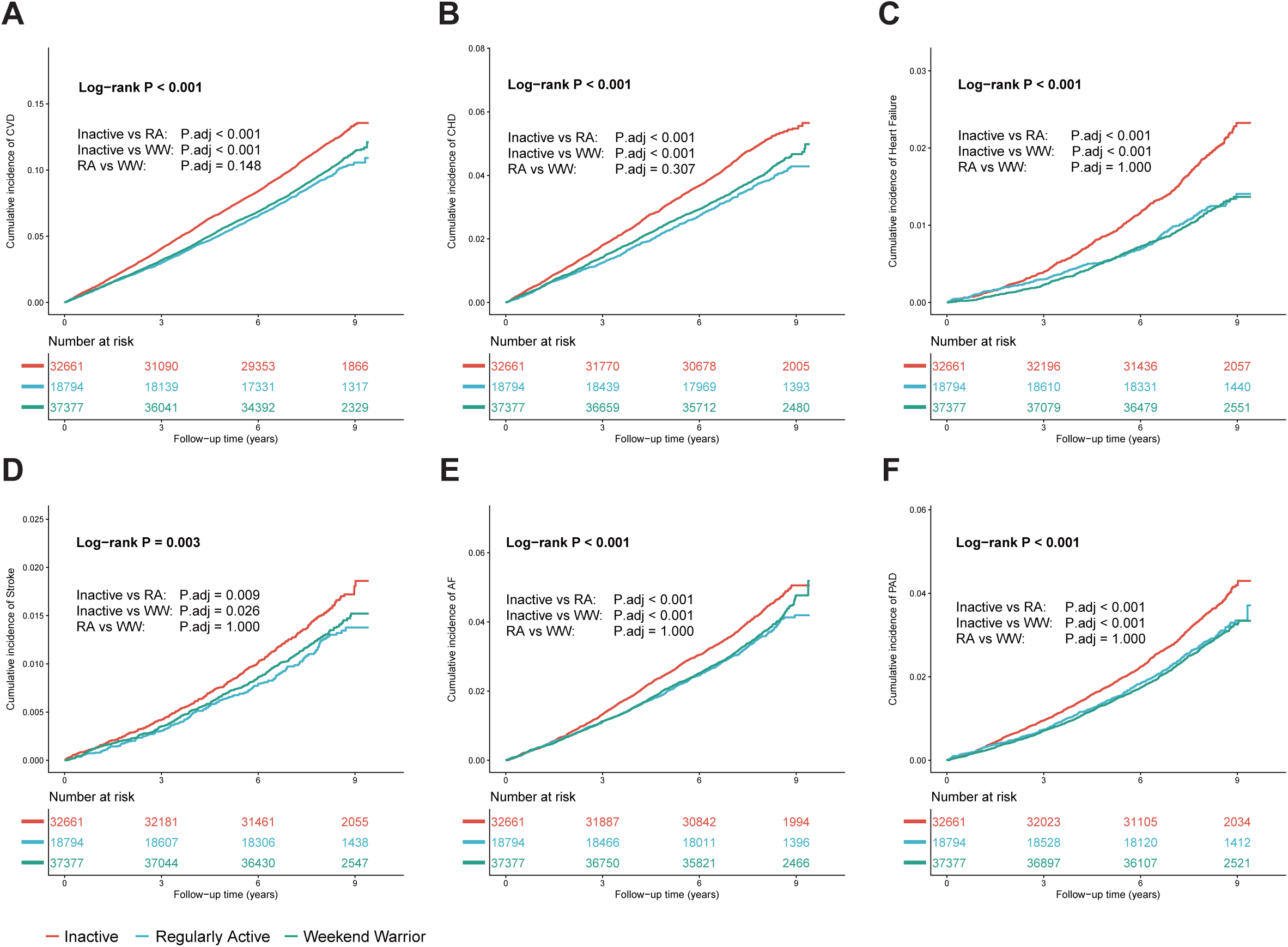
Cumulative incidence of cardiovascular outcomes by physical activity patterns. Kaplan-Meier curves illustrate the cumulative incidence of (A) Total cardiovascular disease (CVD), (B) Coronary heart disease (CHD), (C) Heart failure, (D) Stroke, (E) Atrial fibrillation (AF), and (F) Peripheral artery disease (PAD) among participants with CKM syndrome Stages 0-3. The study population is stratified into three groups: Inactive (red line), Regularly Active (RA; blue line), and Weekend Warrior (WW; green line). The P values represent the overall difference among groups derived from the log-rank test. P.adj indicates the adjusted P value for pairwise comparisons. Abbreviations: AF, atrial fibrillation; CHD, coronary heart disease; CVD, cardiovascular disease; HF, heart failure; PAD, peripheral artery disease; RA, regularly active; WW, weekend warrior.

Given the complex metabolic burden inherent to the CKM syndrome, we performed multivariable Cox regression analyses to control for potential confounders (Table 2). In the fully adjusted model (Model 3), which accounted for BMI, metabolic comorbidities, and medication use, both active patterns were consistently associated with robust risk reductions. Compared with the Inactive group, the RA and WW groups showed similar protection against Total CVD (HR 0.88 and 0.87, respectively; both P < 0.001) and CHD (HR 0.86 for both; both P < 0.001). These findings indicate that achieving guideline-recommended physical activity, regardless of the pattern, confers stable cardiovascular protection across CKM syndrome Stages 0 to 3. For HF, while both patterns were protective, the WW pattern demonstrated a numerically stronger risk reduction (HR 0.72, 95% CI 0.63–0.82, P < 0.001) compared with the RA group (HR 0.81, 95% CI 0.69–0.95, P = 0.010). Comparisons between Model 2 (adjusted for lifestyle but not metabolic mediators) and Model 3 revealed potential mechanistic differences between patterns. For Stroke, significant risk reductions observed in Model 2 for both active groups were attenuated and became non-significant in the RA group (P = 0.167) and borderline in the WW group (P = 0.096) after further adjustment for metabolic risk factors. This suggests that the protective effect of exercise on stroke in this population is largely mediated by improvements in metabolic profiles. Notably, distinct associations were observed for AF and PAD. In the total effect model (Model 2), the RA pattern was significantly associated with a lower risk of AF (HR 0.87, P = 0.003) and showed a trend for PAD (HR 0.91, P = 0.075). However, these associations were fully attenuated in Model 3 (AF: P = 0.444; PAD: P = 0.196), implying that the benefits of regular activity on these outcomes are primarily driven by metabolic improvements. In contrast, the WW pattern retained statistically significant benefits even after full adjustment for metabolic confounders, reducing the risk of AF by 9% (HR 0.91, 95% CI 0.85–0.99, P = 0.024) and PAD by 15% (HR 0.85, 95% CI 0.78–0.93, P < 0.001). This implies that the concentrated, higher-intensity nature of the WW pattern may offer distinct physiological advantages that persist beyond standard metabolic risk factor control.

**Table 2.** Associations of Physical Activity Patterns with Risk of Incident Cardiovascular Disease and Subtypes in Individuals with CKM Syndrome (Stages 0-3).

| Outcome | PA Pattern | Cases / Total | IR<br>(1000PY) | Model 1 |  | Model 2 |  | Model 3 |  |
| --- | --- | --- | --- | --- | --- | --- | --- | --- | --- |
|  |  |  |  | HR (95% CI) | P Value | HR (95% CI) | P Value | HR (95% CI) | P Value |
| **Total CVD** | Inactive | 3,778 / 32,661 | 15.51 | 1.00 [Reference] |  | 1.00 [Reference] |  | 1.00 [Reference] |  |
|  | Regularly Active | 1,729 / 18,794 | 12.07 | 0.78 (0.73–0.82) | < 0.001 | 0.82 (0.77–0.86) | < 0.001 | 0.88 (0.83–0.94) | < 0.001 |
|  | Weekend Warrior | 3,618 / 37,377 | 12.77 | 0.77 (0.74–0.81) | < 0.001 | 0.81 (0.77–0.85) | < 0.001 | 0.87 (0.83–0.91) | < 0.001 |
| **CHD** | Inactive | 1,611 / 32,661 | 6.42 | 1.00 [Reference] |  | 1.00 [Reference] |  | 1.00 [Reference] |  |
|  | Regularly Active | 706 / 18,794 | 4.82 | 0.71 (0.65–0.78) | < 0.001 | 0.78 (0.73–0.84) | < 0.001 | 0.86 (0.78–0.94) | < 0.001 |
|  | Weekend Warrior | 1,505 / 37,377 | 5.18 | 0.73 (0.68–0.78) | < 0.001 | 0.78 (0.73–0.84) | < 0.001 | 0.86 (0.80–0.93) | < 0.001 |
| **HF** | Inactive | 596 / 32,661 | 2.34 | 1.00 [Reference] |  | 1.00 [Reference] |  | 1.00 [Reference] |  |
|  | Regularly Active | 219 / 18,794 | 1.47 | 0.82 (0.70–0.96) | 0.013 | 0.71 (0.60–0.83) | < 0.001 | 0.81 (0.69–0.95) | 0.010 |
|  | Weekend Warrior | 418 / 37,377 | 1.42 | 0.82 (0.72–0.93) | 0.002 | 0.63 (0.56–0.72) | < 0.001 | 0.72 (0.63–0.82) | < 0.001 |
| **Stroke** | Inactive | 486 / 32,661 | 1.91 | 1.00 [Reference] |  | 1.00 [Reference] |  | 1.00 [Reference] |  |
|  | Regularly Active | 223 / 18,794 | 1.5 | 0.85 (0.77–0.93) | < 0.001 | 0.86 (0.73–1.01) | 0.061 | 0.89 (0.76–1.05) | 0.167 |
|  | Weekend Warrior | 474 / 37,377 | 1.61 | 0.81 (0.75–0.87) | < 0.001 | 0.86 (0.76–0.98) | 0.024 | 0.89 (0.78–1.02) | 0.096 |
| **AF** | Inactive | 1,389 / 32,661 | 5.52 | 1.00 [Reference] |  | 1.00 [Reference] |  | 1.00 [Reference] |  |
|  | Regularly Active | 672 / 18,794 | 4.58 | 0.65 (0.55–0.76) | < 0.001 | 0.87 (0.79–0.95) | 0.003 | 0.96 (0.88–1.06) | 0.444 |
|  | Weekend Warrior | 1,386 / 37,377 | 4.76 | 0.57 (0.50–0.65) | < 0.001 | 0.83 (0.77–0.90) | < 0.001 | 0.91 (0.85–0.99) | 0.024 |
| **PAD** | Inactive | 1,110 / 32,661 | 4.38 | 1.00 [Reference] |  | 1.00 [Reference] |  | 1.00 [Reference] |  |
|  | Regularly Active | 526 / 18,794 | 3.57 | 0.85 (0.77–0.95) | 0.003 | 0.91 (0.82–1.01) | 0.075 | 0.93 (0.84–1.04) | 0.196 |
|  | Weekend Warrior | 1,010 / 37,377 | 3.45 | 0.77 (0.70–0.84) | < 0.001 | 0.83 (0.76–0.90) | < 0.001 | 0.85 (0.78–0.93) | < 0.001 |

Note: Data are presented as Hazard Ratios (HR) and 95% Confidence Intervals (CI). The Inactive group served as the reference group. Incidence Rate (IR) is expressed per 1,000 person-years. Model 1: Adjusted for age, sex, and ethnicity. Model 2: Adjusted for Model 1 covariates plus Townsend Deprivation Index (TDI), education level, smoking status, alcohol consumption, Healthy Diet Score (HDS), Sleep Score, and estimated glomerular filtration rate (eGFR<sub>cr-cys</sub>). Model 3: Further adjusted for body mass index (BMI), baseline clinical history (hypertension, diabetes, dyslipidemia), and medication use (including antihypertensives, lipid-lowering agents, and insulin or oral hypoglycemics). Abbreviations: AF, atrial fibrillation; CHD, coronary heart disease; CKM, cardiovascular-kidney-metabolic; CVD, cardiovascular disease; HF, heart failure; PAD, peripheral artery disease; PA, physical activity; PY, person-years.

### 3.3 Stratified Analysis by CKM Stages

To evaluate the stability of physical activity benefits across disease progression, we performed stratified analyses by CKM stages. In the fully adjusted model (Model 3), no significant interactions were observed between physical activity patterns and CKM stages for any outcome (all P for interaction> 0.3) (Fig. 2, Fig. S2), indicating generally consistent protective trends across the CKM spectrum. For the primary outcome (total CVD), significant risk reductions were observed in the large population with established metabolic risk factors (Stage 2), where both RA (HR 0.90, P = 0.001) and WW (HR 0.88, P < 0.001) patterns were effective. Most notably, in participants with advanced disease (Stage 3), the protective effects were profound. The WW pattern was associated with a 38% reduction in CVD risk (HR 0.62, 95% CI 0.47–0.83, P = 0.001), and the RA pattern showed a similar benefit (HR 0.58, 95% CI 0.40–0.86, P = 0.006) (Fig. 2). Results for CHD generally mirrored these findings, with both patterns showing significance in Stage 2. In Stage 3, significant benefit was observed for the RA pattern (P = 0.017), while the WW pattern showed a borderline association (P = 0.056) (Fig. S2A). Intriguingly, for HF and PAD, the WW pattern demonstrated unique efficacy in advanced stages. In Stage 3 participants, the WW pattern was significantly associated with a marked reduction in HF risk (HR 0.47, 95% CI 0.24–0.90, P = 0.022) and PAD risk (HR 0.52, 95% CI 0.31–0.89, P = 0.019). In contrast, the RA group did not achieve statistical significance for these outcomes in Stage 3 (HF: P = 0.49; PAD: P = 0.10), suggesting that concentrated physical activity might be particularly advantageous for vascular and cardiac pump function in patients with severe CKM progression (Fig. S2B, F). For Stroke and AF, stratified analyses did not reveal consistent significant associations across stages in the fully adjusted model, likely due to the limited number of events in subgroups and the strong mediating role of metabolic control for these outcomes (Fig. S2C, D).

**Figure 2.**
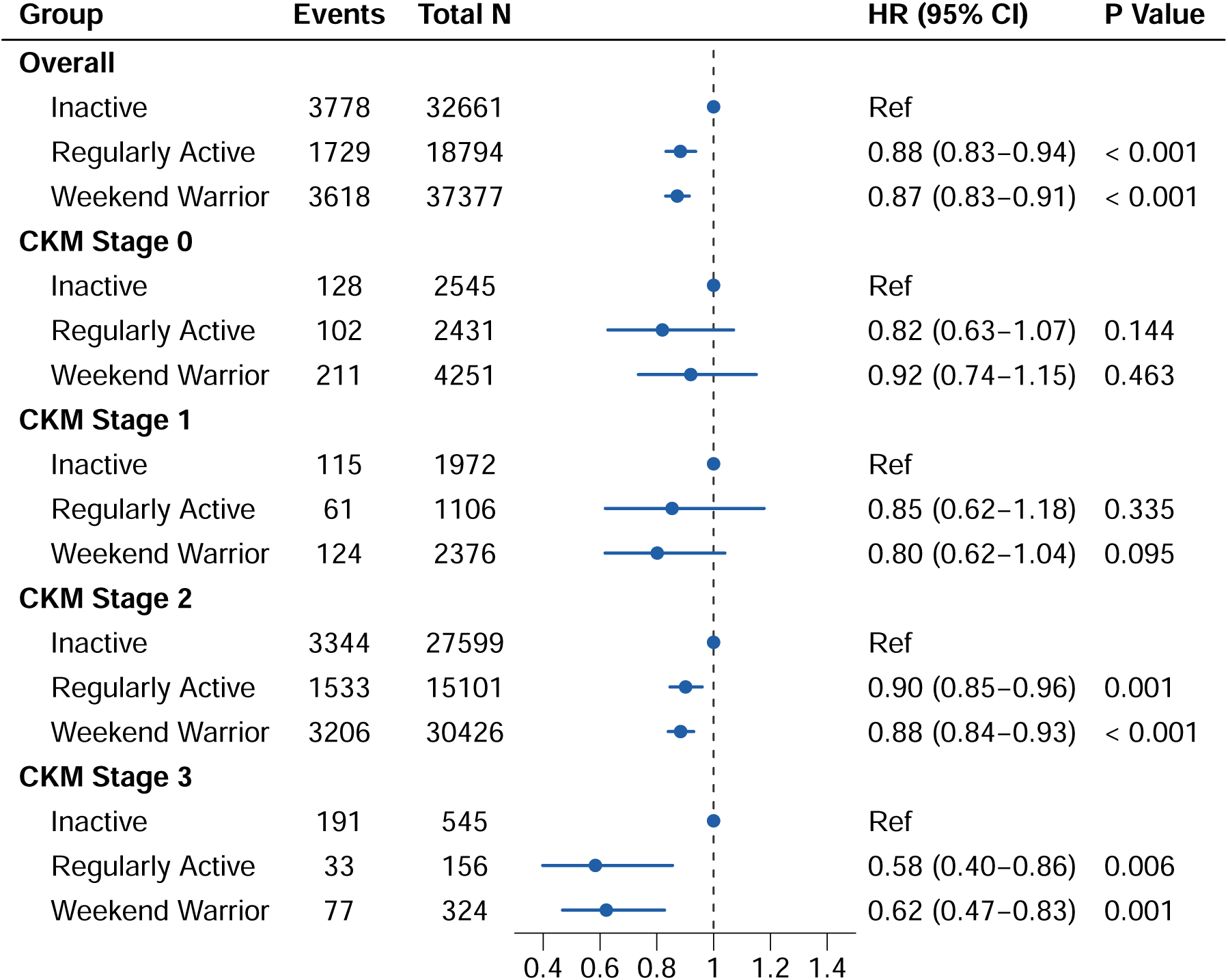
Association of physical activity patterns with the risk of total cardiovascular disease stratified by CKM stages. Forest plot displaying the Hazard Ratios (HRs) and 95% Confidence Intervals (CIs) for incident total CVD across the overall population and subgroups stratified by CKM stages (0 to 3). The Inactive group serves as the reference category (Ref). Results are derived from the fully adjusted multivariable Cox proportional hazards model (Model 3). P for interaction = 0.349 across CKM stages. Abbreviations: CI, confidence interval; CKM, cardiovascular-kidney-metabolic; CVD, cardiovascular disease; HR, hazard ratio; Ref, reference.

### 3.4 Dose-Response Associations of Physical Activity with Cardiovascular Outcomes across CKM Stages

We utilized restricted cubic splines to characterize the dose-response association between MVPA volume and cardiovascular outcomes. For total CVD, we observed a curvilinear, L-shaped association (P non-linearity < 0.001, Fig. 3A). The risk of CVD decreased steeply with increasing MVPA volume up to approximately 200–300 minutes/week, after which the protective effect plateaued. This suggests that the most substantial marginal benefits are gained by moving from inactivity to meeting the lower bounds of guideline-recommended levels. Similar non-linear patterns with steep initial risk reductions were observed for CHD, HF, and PAD (all P for non-linearity <0.05) (Fig. 3B, C, F). Analyses using the full range of MVPA data yielded consistent results (Fig. S3).

**Figure 3.**
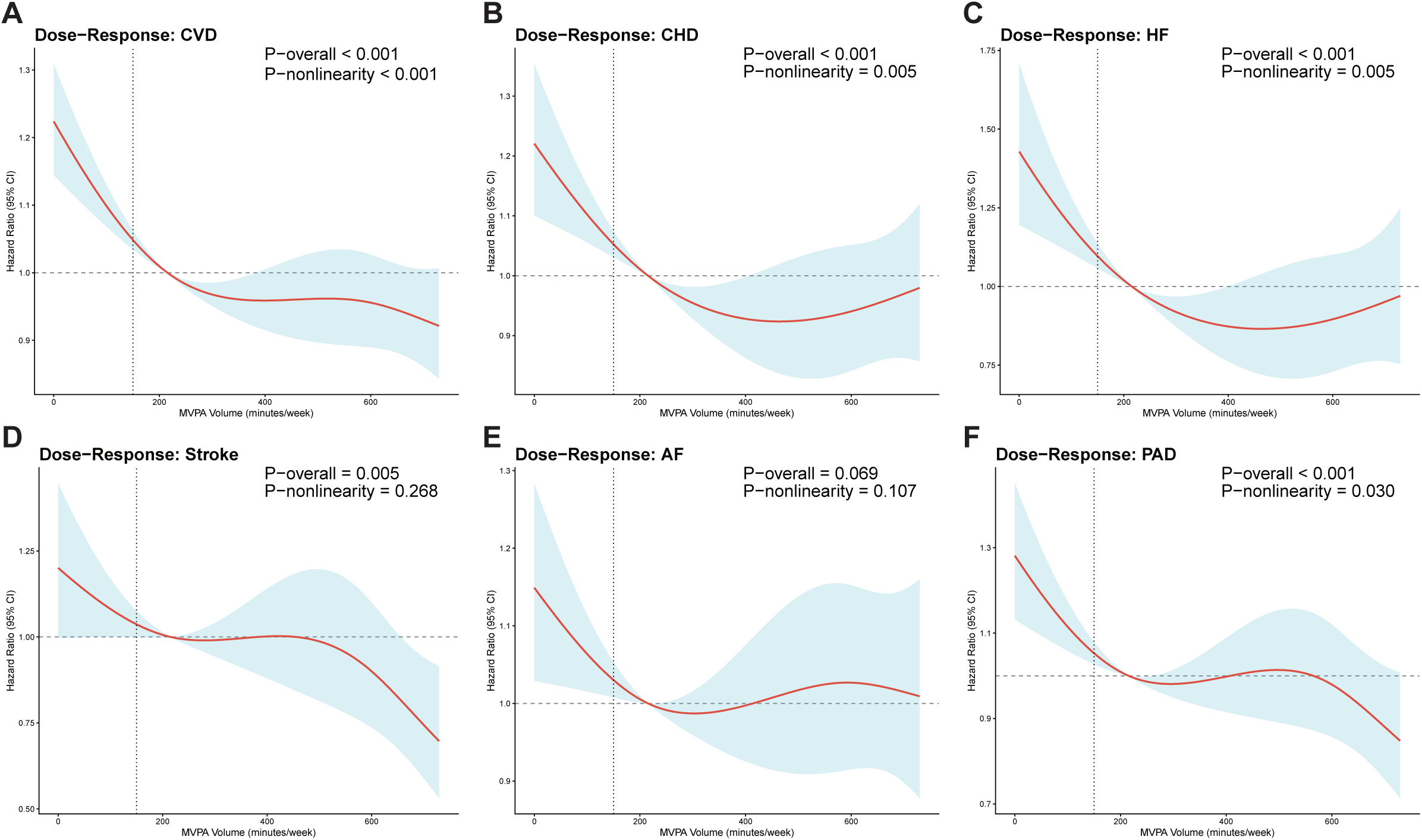
Dose-response associations of moderate-to-vigorous physical activity (MVPA) volume with incident cardiovascular outcomes. Restricted cubic splines illustrating the associations between weekly MVPA volume (minutes/week) and the risk of (A) Total CVD, (B) CHD, (C) Heart Failure, (D) Stroke, (E) Atrial Fibrillation, and (F) PAD. Models were fitted with four knots placed at the 5th, 35th, 65th, and 95th percentiles. Continuous MVPA values were winsorized at the 95th percentile to minimize the influence of extreme values. The solid red line represents the estimated Hazard Ratio (HR) derived from the fully adjusted model (Model 3), and the blue vshaded area indicates the 95% Confidence Interval (CI). The horizontal dashed line represents the reference hazard ratio of 1.0, and the vertical dashed line represents the guideline-recommended level of 150 min/week. Abbreviations: CHD, coronary heart disease; CI, confidence interval; CVD, cardiovascular disease; HF, heart failure; HR, hazard ratio; MVPA, moderate-to-vigorous physical activity; PAD, peripheral artery disease.

To explore whether the optimal dose of physical activity varies with disease severity, we performed analyses stratified by CKM stages (Fig. S4, Table S4). In participants with established metabolic risk factors (CKM Stage 2), which accounted for the majority of recorded events (N=8,083 for total CVD), we observed strong evidence of non-linear inverse associations (Fig. S4A). Specifically, for total CVD (P for overall < 0.001; P for non-linearity < 0.001), CHD (P for non-linearity = 0.006), and HF (P for non-linearity = 0.004), the curves displayed a distinct L-shaped pattern (Fig. S4A-C). A steep reduction in risk was observed at lower levels of physical activity, which then plateaued at higher volumes, suggesting that the greatest marginal benefit is gained by transitioning from inactivity to moderate activity. Interestingly, Stroke in Stage 2 exhibited a linear inverse association (P for overall = 0.012; P for non-linearity = 0.740), indicating a continuous risk reduction with increasing activity volume without a clear plateau (Fig. S4D).

In the advanced disease group (CKM Stage 3), despite a smaller sample size, significant protective associations persisted for total CVD (P for overall = 0.001), CHD (P for overall = 0.016) and PAD (P for overall = 0.044) (Fig. S4A, B, F). Notably, the curve for CVD in Stage 3 maintained a non-linear shape (P for non-linearity = 0.029), reinforcing the benefits of physical activity even in severe disease states (Fig. S4A). Conversely, in the early stages (CKM Stages 0 and 1), no statistically significant associations were observed across outcomes (P for overall > 0.05), likely attributable to the lower event rates and lower baseline cardiovascular risk in these subgroups (Fig. S4A-F).

### 3.5 Joint Association of Physical Activity and Systemic Inflammation with Cardiovascular Outcomes

Given that chronic systemic inflammation is a central pathophysiological driver of CVD, we selected hs-CRP as the representative biomarker to investigate whether the cardioprotective effects of PA are modulated by systemic inflammatory burden [22]. Subgroup analyses stratified by CKM stages demonstrated that the cardiovascular benefits of PA were generally consistent regardless of systemic inflammation status. In individuals with low-to-moderate inflammation (hs-CRP ≤ 3 mg/L), both PA patterns were associated with reduced CVD risk (Fig. 4A, Table S5). Notably, in Stage 2 participants, both WW (HR 0.90, 95% CI 0.85–0.95, P < 0.001) and RA patterns (HR 0.92, 95% CI 0.86–0.99, P = 0.020) significantly lowered CVD risk compared to the inactive group. Similarly, in Stage 3 participants with non-elevated hs-CRP, substantial risk reductions were observed for both RA (HR 0.57, 95% CI 0.35–0.92, P = 0.023) and WW patterns (HR 0.67, 95% CI 0.47–0.95, P = 0.025). In the elevated hs-CRP subgroup, PA continued to exhibit protective effects, particularly in advanced CKM stages. For participants in Stage 2 with elevated hs-CRP, the WW pattern was associated with a significant 15% reduction in CVD risk (HR 0.85, 95% CI 0.76–0.96, P = 0.007). Most strikingly, among the highest-risk group (Stage 3 with elevated hs-CRP), the WW pattern showed a profound CVD risk reduction of 54% (HR 0.46, 95% CI 0.25–0.84, P = 0.012), while the RA pattern also demonstrated a protective trend (HR 0.48, 95% CI 0.23–1.01, P = 0.053).

**Figure 4.**
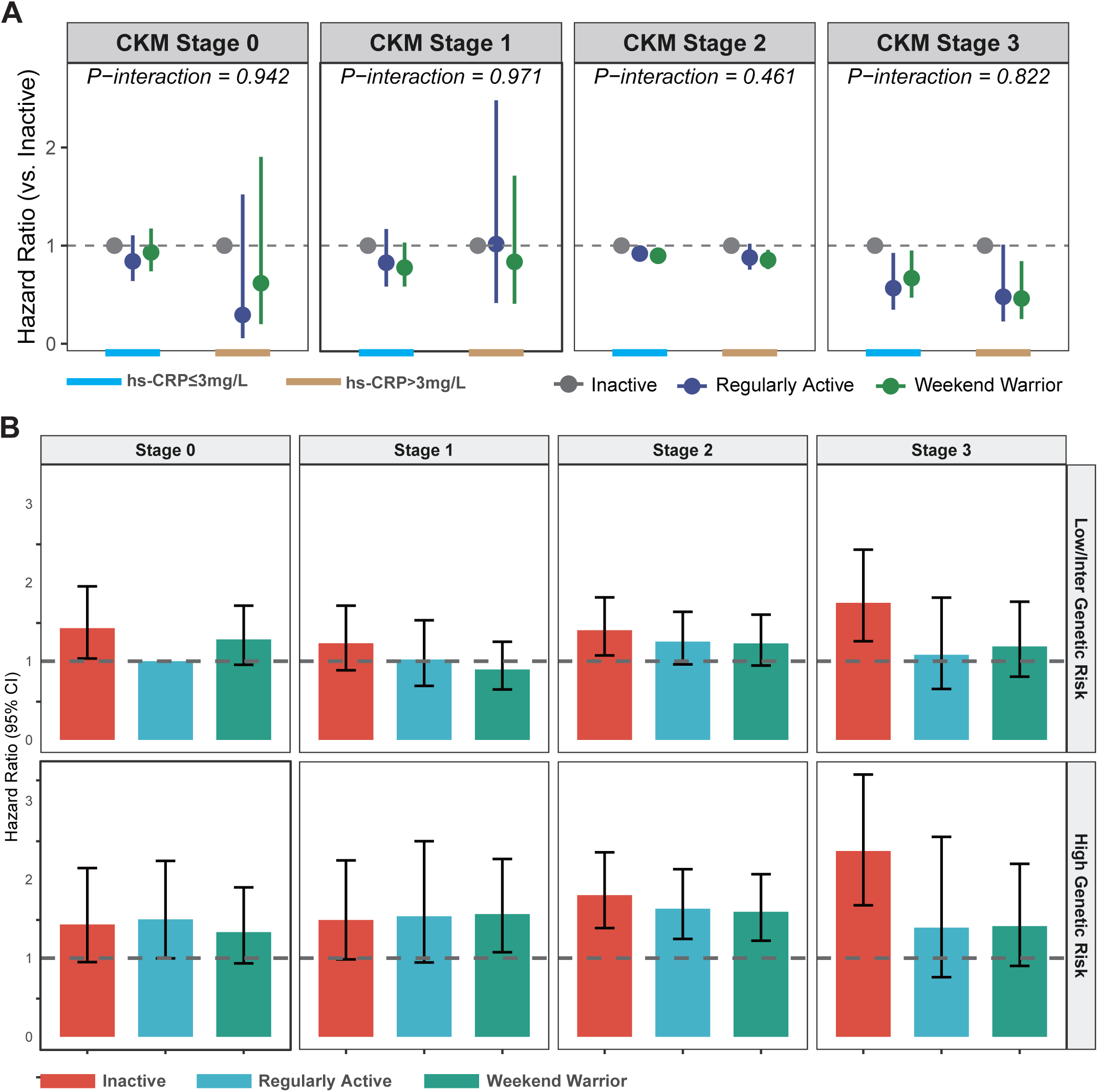
Joint associations of physical activity with systemic inflammation and genetic susceptibility on cardiovascular risk across CKM stages. (A) Hazard Ratios for total CVD stratified by systemic inflammation status (high-sensitivity C-reactive protein [hs-CRP] levels) across CKM Stages 0-3. The Inactive group serves as the reference (HR = 1.0) within each subgroup. The forest plot compares Regularly Active (blue) and Weekend Warrior (green) patterns against the Inactive group (grey) in participants with low-to-moderate (≤3 mg/L) and elevated (> 3 mg/L) hs-CRP. Abbreviations: CI, confidence interval; CKM, cardiovascular-kidney-metabolic; CVD, cardiovascular disease; HR, hazard ratio; hs-CRP, high-sensitivity C-reactive protein. (B) Joint association of physical activity patterns and genetic susceptibility (Polygenic Risk Score for CHD) with CVD risk across CKM Stages 0-3. The group “Stage 0 + Low/Intermediate Genetic Risk + Regularly Active” serves as the single global reference (indicated by the dashed line at HR = 1.0). Bars represent the HRs and 95% CIs for Inactive (red), Regularly Active (blue), and Weekend Warrior (green) participants stratified by CKM stages and genetic risk category (Low/Intermediate vs. High).

The interaction tests between PA patterns and systemic inflammatory status were non-significant across all CKM stages (all P-interaction > 0.4). These findings suggest that the cardiovascular benefits of physical activity are not attenuated by systemic inflammation. Even in individuals with advanced CKM syndrome and high inflammatory burden, engagement in physical activity—particularly the “Weekend Warrior” pattern—remains a highly effective strategy for cardiovascular protection.

### 3.6 Joint Association of Physical Activity and Genetic Susceptibility with Cardiovascular Outcomes across CKM Stages

To evaluate whether physical activity could counteract the congenital genetic risk in patients across the CKM spectrum, we utilized the PRS for coronary heart disease to conduct a joint analysis (Fig. 4B, Table S6).

First, we observed no significant multiplicative interaction between physical activity patterns and genetic risk categories in the overall population (P for interaction = 0.894). This indicates that the relative cardiovascular benefits of physical activity are generally consistent across different genetic backgrounds. However, further joint analyses using the “Stage 0 + low/intermediate PRS + Regularly Active” group as the single reference (HR = 1.00) revealed a significant offsetting effect of physical activity against genetic susceptibility.

In the Stage 0 (healthy) population, physical inactivity emerged as a risk factor comparable to high genetic predisposition. Individuals with low/intermediate genetic risk who were inactive had a significantly increased CVD risk compared to the active reference group (HR 1.42, 95% CI 1.03–1.95, P = 0.031). Notably, for individuals with high genetic risk, engaging in physical activity attenuated the risk to levels similar to inactive low/intermediate-risk individuals. The Hazard Ratios for the RA group (HR 1.49, 95% CI 1.00–2.23, P = 0.052) and the WW group (HR 1.33, 95% CI 0.93–1.90, P = 0.116) were statistically comparable to the risk conferred by inactivity alone in the low-risk group. In the Stage 2 population, which carries a heavier metabolic burden, the combination of high genetic risk and inactivity led to the highest risk (HR 1.80, 95% CI 1.38–2.34, P < 0.001). However, maintaining an active lifestyle was associated with lower risk estimates. In the high-risk group, both the RA (HR 1.63, 95% CI 1.24–2.13, P < 0.001) and WW pattern (HR 1.59, 95% CI 1.22– 2.07, P = 0.001) showed reduced hazard ratios compared to the inactive group. This demonstrates that even when facing the double hit of genetic susceptibility and established metabolic risk factors, adhering to guideline-recommended activity levels remains protective. The most pronounced difference in point estimates was observed in Stage 3. High-genetic-risk individuals who were inactive faced a greater than 2-fold risk increase (HR 2.36, 95% CI 1.67–3.33, P < 0.001). In contrast, those who were active exhibited substantially attenuated risk estimates. Although the associations did not reach statistical significance—likely due to the limited sample size in this advanced-stage subgroup—the magnitude of the reduction was clinically relevant (RA: HR 1.39, 95% CI 0.76–2.54, P = 0.289; WW: HR 1.41, 95% CI 0.90–2.20, P = 0.134). These findings suggest a clinically meaningful potential for physical activity to blunt the impact of high genetic risk even in advanced disease stages.

### 3.7 Sensitivity and Subgroup Analyses

The cardiovascular benefits of physical activity patterns demonstrated high robustness across a series of sensitivity analyses (Fig. S5-9). First, regarding the definition of physical activity volume, protective associations persisted regardless of the MVPA thresholds used. Most notably, when lowering the active threshold to 75 min/week (approx. 25th percentile; Fig. S5), we observed substantial risk reductions for both WW (HR 0.83, 95% CI 0.79–0.88, P < 0.001) and RA groups (HR 0.82, 95% CI 0.77–0.88, P < 0.001). This suggests that in the CKM population, accumulating even half the guideline-recommended activity level confers significant cardiovascular protection compared with inactivity. Conversely, when the reference group was expanded to include individuals active up to the 75th percentile (381 min/week; Fig. S7), the hazard ratios attenuated slightly but remained statistically significant (WW: HR 0.91, P = 0.004; RA: HR 0.88, P < 0.001), as expected due to the healthier reference comparator. Second, applying a stricter definition for the WW pattern (≥75% of weekly MVPA in 1–2 days; Fig. S8) yielded results consistent with the main analysis (WW: HR 0.86, 95% CI 0.79–0.93, P < 0.001). This confirms that the observed benefits are attributable to the concentrated activity pattern itself rather than spillover activity distributed across the week. Third, in the landmark analysis excluding CVD events within the first 2 years to minimize reverse causality (Fig. S9), risk estimates were materially unchanged (WW: HR 0.88, P < 0.001; RA: HR 0.88, P < 0.001). Furthermore, when employing the Fine-Gray model to account for the competing risk of non-cardiovascular death—a critical consideration in multimorbid CKM participants—the associations remained significant and similar in magnitude to the standard Cox analysis (WW: HR 0.88, 95% CI 0.84–0.92, P < 0.001; RA: HR 0.89, 95% CI 0.84–0.94, P < 0.001) (Table S7). Finally, to assess the overall survival benefit, we evaluated all-cause mortality. Both the WW (HR 0.76, 95% CI: 0.70–0.81, P < 0.001) and RA (HR: 0.79, 95% CI: 0.73–0.87, P < 0.001) patterns demonstrated robust protection against death from any cause, reinforcing the comprehensive health benefits of achieving recommended physical activity levels across CKM stages (Table S7).

In subgroup analyses (Fig. S10), the associations were generally consistent across most subgroups. However, significant interactions were observed for sex (P for interaction = 0.004) and diabetes status (P for interaction = 0.042). The protective magnitude of the WW pattern appeared stronger in women (HR 0.81 vs. 0.93 in men) and in participants with diabetes (HR 0.73 vs. 0.88 in non-diabetics), highlighting the particular importance of physical activity in these groups.

## 4. Discussion

This large-scale prospective study, involving over 80,000 UK Biobank participants, is the first to systematically evaluate the association between accelerometer-defined physical activity patterns and cardiovascular outcomes across the different stages of CKM syndrome (Stages 0–3). Our main findings are as follows: First, across CKM stages, the WW pattern and the RA pattern exhibited similar protective effects in reducing the risk of total CVD, independent of traditional metabolic risk factors. Second, the protective effect of physical activity is highly universal; even in high-risk individuals in advanced stages of disease progression (Stage 3), or those with high systemic inflammation or high genetic susceptibility, increasing activity levels significantly offsets risk. and notably, regarding specific subtypes such as HF, PAD, and AF, concentrated physical activity (WW) showed a more robust independent association than regular activity in fully adjusted models. These findings fill a critical evidence gap regarding the effectiveness of exercise patterns in CKM syndrome management, affirming that achieving targets for total volume and intensity is the core priority within complex metabolic contexts.

Our research builds upon the landmark work of Khurshid et al. regarding the general population [7], extending the scope into the specific clinical evolution of CKM syndrome. We excluded individuals with a baseline diagnosis of CVD (Stage 4) to focus on primary prevention during CKM progression, confirming the robust protective effects of PA patterns across CKM risk strata. While the foundational research by Khurshid et al. established key benchmarks for the general population, patients with CKM syndrome present with a more complex metabolic profile. Consequently, we extended their methodology by applying more stringent adjustment models—accounting for BMI, baseline comorbidities, and medication—to determine if the observed protective effect is independent of conventional metabolic pathways. Consistent with prior research [6], we found that both activity patterns reduce CVD risk.

However, a key divergence emerged in subtype analyses. In our fully adjusted model, the associations for the RA group with Stroke, AF, and PAD were attenuated to non-significance, whereas the WW pattern maintained a robust risk reduction for HF, AF, and PAD. This discrepancy suggests that the benefits of regular, moderate-intensity activity may be largely mediated through weight management and metabolic control [23], while the WW pattern may provide additional vascular benefits through alternative pathways. A plausible explanation lies in the intensity hypothesis, which posits that WW exercise typically involves higher-intensity, longer-duration bouts of exertion. Vigorous physical activity is known to be superior to low-to-moderate intensity exercise in improving cardiorespiratory fitness (VO_2_max), endothelial function, and vascular remodeling [24,25]. These factors are critical determinants of risk for HF and PAD—outcomes that are particularly sensitive to improvements in arterial stiffness and cardiac pump function [26–28]—and AF, which is influenced by autonomic tone and atrial remodeling. Crucially, the significant reduction in all-cause mortality demonstrates that this concentrated, high-intensity pattern is not only effective but also safe, conferring survival benefits comparable to daily exercise even in high-risk CKM populations.

Importantly, the protective effect of physical activity did not diminish as CKM stages progressed, remaining statistically significant even in Stage 3 patients. In this advanced stage, high metabolic derangement and chronic systemic inflammation often lead clinicians to rely heavily on pharmacological interventions, potentially overlooking the power of lifestyle modification. However, our study confirms that the benefits of physical activity were neither weakened by elevated systemic inflammation nor offset by high genetic risk. This suggests that in late-stage CKM syndrome, exercise may operate through non-metabolic pathways, such as direct anti-inflammatory effects [29,30], suppression of oxidative stress, improved clearance of uremic toxins, and enhanced microcirculatory function [31]. Our findings underscore the need for a full-course intervention philosophy in CKM management; physical activity should not only be a preventive tool in early stages but also a core component of late-stage risk management.

Notably, subgroup analyses revealed that women and patients with diabetes derived greater benefits from the WW pattern, offering significant clinical guidance. For diabetic patients, high-intensity concentrated exercise may more effectively induce GLUT4 translocation in skeletal muscle, leading to more sustained insulin sensitization [32]. Regarding sex-specific differences, women may exhibit superior vascular adaptive responses to the same dose of physical activity compared to men, aligning with recent research on sex-specific exercise physiology [33]. In a related study, Ji et al. [34] further highlighted that while women may have a lower prevalence of CKM Stage 3, they experience disproportionately higher excess mortality risks across the CKM spectrum, underscoring the critical importance of high-intensity physical activity interventions in this demographic. These findings reinforce the status of “Exercise as Medicine” in CKM syndrome management: even for patients with complex conditions, high inflammatory burdens, or metabolic disorders, persistent activity can reverse adverse prognoses. In contrast, the non-significant results observed in Stages 0 and 1 likely reflect a statistical power issue due to the low baseline risk and limited number of events, rather than a lack of biological efficacy. Long-term protective effects in these early groups may require more extended follow-up to manifest.

### Limitations

First, despite multivariate adjustment and sensitivity analyses, residual confounding cannot be entirely ruled out, and a definitive causal relationship between PA and CV benefits cannot be established. Second, PA data were captured via a 7-day accelerometer at baseline, which may not reflect changes in exercise habits over the multi-year follow-up period. However, adult PA patterns are generally stable, and accelerometry avoids the recall bias inherent in questionnaires. Third, while we excluded events occurring within the first two years of follow-up to minimize reverse causation (where illness leads to reduced activity), this bias cannot be completely eliminated. Fourth, the data collection predated the widespread clinical use of SGLT2 inhibitors and GLP-1 receptor agonists, which are now foundational for CKM syndrome management [35,36]. While our analysis adjusted for conventional glucose-lowering and antihypertensive medications, potential synergistic effects between these drugs and physical activity could not be explored. Last, the UK Biobank cohort is predominantly white and healthier than the general UK population, which may limit the extrapolability of these findings to other ethnicities or lower socioeconomic groups.

## Conclusion

Our findings underscore that the WW physical activity pattern is associated with a significant reduction in cardiovascular disease risk. This protective effect is universal across CKM Stages 0–3, remaining robust even among those with systemic inflammation, or those with high genetic susceptibility. Furthermore, the WW pattern may offer unique, independent advantages in mitigating the risks of HF, AF, and PAD, with particularly pronounced benefits observed in women and patients with diabetes. These findings carry substantial public health and clinical implications: for CKM patients who face barriers to daily exercise, concentrating high-quality physical activity into one or two days is a feasible and highly effective cardiovascular protection strategy. Clinicians should adopt an “every move counts” philosophy when prescribing lifestyle interventions, emphasizing total volume and intensity over frequency. To achieve optimal cardiovascular benefits, we recommend a weekly target of 200–300 minutes of MVPA.

## Data availability

The datasets analyzed during the current study are available from the UK Biobank (Application ID 545732). As a large-scale biomedical repository, access to the UK Biobank data is subject to a formal application and approval process via the official website (https://www.ukbiobank.ac.uk/use-our-data/apply-for-access).

## Ethics approval and consent to participate

The study protocols were approved by the North West Multi-centre Research Ethics Committee (REC reference: 21/NW/0157) and the UK National Information Governance Board. All participants gave their written informed consent when enrolled in the UK Biobank.

## Consent for publication

The submission of this manuscript has been approved by all authors, who have read and consented to the publication of the final version.

## Acknowledgements

We are grateful to the UK Biobank (Application ID 545732) for providing the data essential to this work. Our sincere thanks go to the participants who contributed their information to this resource, as well as the staff and management teams whose efforts made this study possible.

